# Respiratory syncytial virus infection and reinfection patterns during a community outbreak in Kenya investigated by whole genome sequencing, 2023/2024

**DOI:** 10.64898/2026.01.26.26343753

**Authors:** Edidah M. Ong’era, Esther. N. Katama, Joyce U. Nyiro, Arnold W. Lambisia, John M. Morobe, Nickson Murunga, Samuel Mwasya, Martin Mutunga, Clement Lewa, George Githinji, Philip Bejon, Charles J. Sande, E. Wangeci Kagucia, Simon Delicour, D. James Nokes, Patrick K. Munywoki, Edward C Holmes, Charles N. Agoti

## Abstract

**Background:** Respiratory syncytial virus (RSV) is a leading cause of severe acute respiratory infection in infants, young children and vulnerable adults. Despite implications for designing interventions, our understanding of RSV infection/reinfection patterns during community outbreaks is incomplete.

**Methods:** To characterize respiratory virus infections regardless of symptom status, we performed a prospective cohort surveillance in coastal Kenya from August 2023 to August 2024. Nasopharyngeal/oropharyngeal (NP/OP) swabs were collected 1-2 times weekly regardless of symptom status for quantitative PCR testing followed by genomic analysis. RSV reinfections were defined as two positive tests separated by ≥14 days and with intervening ≥1 negative tests.

**Results:** Of 672 individuals screened, 74 tested positive (93/22,000 swabs; 0.4%). The median age among infections was 4.2 years (interquartile range (IQR): 1.8-9.4), 58.1% being female versus a median age of 14.3 years (IQR: 4.8-29.6) and 64.4% female among uninfected individuals. Overall incidence rate was 19.8 infections/100 person-years, highest incidence among infants (174.0/100 person-years, 95% CI:103.0-274.0). Infection episodes fell into seven viral lineages: A.D (n=1), A.D.1 (n=2), A.D.1.11 (n=21), A.D.2.1 (n=19), A.D.3 (n=30), A.D.5.2 (n=1), and B.D.E.1 (n=2). Six individuals (8.1%; 13.7/100 person-years) experienced reinfections, three involving same viruses with 0-3 nucleotide differences across the entire RSV genome, while other three had 20,78 and 200 nucleotide differences. The (suspect) reinfected individuals were all under 2 years of age, included both males and females, and had no reported chronic illnesses.

**Conclusion:** RSV community infections predominantly occur in children regardless of clinical presentation. Reinfections within the same season are rare.

**Key points:**

- In a community cohort prospective study in coastal Kenya, RSV-A predominated the 2023/24 epidemic and seven lineages co-circulated.
- Overall incidence was 19.6 infections/100 person-years and highest in infants.
- Most reinfections (5/6) were asymptomatic and only half had amino acid changes.

## Main text

Human respiratory syncytial virus (RSV) is a leading cause of hospitalization globally due to acute lower respiratory tract infection (LRTI) in infants and young children [1–4]. In 2019 alone, prior to vaccine licensure, ∼3.6 million children aged under 5 years were hospitalized due to RSV-associated disease resulting in ∼ 100,000 deaths [5,6]. RSV infection in vulnerable adults and elderly populations has also been associated with a disease burden comparable with that of seasonal influenza [6,7]. RSV infections start early in life, and by the age of 3 years all individuals are expected to have been exposed [8,9]. Disease severity is most pronounced in infants, and in frail older adults, whereas most infections in older children and healthy adults are mild and self-limiting [10–12].

RSV reinfections occur at all ages, previously observed to be highest in infants and children < 5 years (∼40/100 person-years), followed by a progressively fall with increasing age before rising again in older adults (>60 years) [13–15]. The occurrence of repeat RSV infections has important implications for the effectiveness of RSV prophylactic interventions [12–14]. Further, repeat infections are perceived to be key to RSV epidemiological persistence in human population and in vulnerable groups can result in disease of comparable severity to that associated with primary infection [14,15]. Repeat RSV infections may occur due to (a) host-related factors (e.g., inadequate/waning immune responses), (b) virus-related factors (e.g., antigenic variation/variability), or (c) environment-related factors (e.g., household crowding) [18–20]. The contribution of each of these factors remains understudied.

RSV sequences can be divided into two genetically and antigenically distinct subgroups (RSVA and RSVB) each containing multiple genetic lineages [21]. An RSV classification system below subgroup level based on phylogenetic analysis of whole genomes proposes that RSV sequences be classified into 40 lineages (24 within RSVA and 16 within RSVB)[22]. The RSV genome comprises a single stranded, non-segmented RNA molecule of ∼15.2 kb length that encodes 11 viral proteins. The two surface proteins - the attachment (G) glycoprotein and the fusion (F) protein - are key in triggering the RSV adaptive humoral immune responses [21,23,24]. The F protein exhibits high sequence conservation across subgroups and lineages, but the G glycoprotein distinguishes between subgroup RSVA and RSVB sequences (with up to 47% amino acid sequence divergence) [11–13].

Although considerable work has been done on RSV reinfections, especially in the <5 years-old age group in a hospital setting, few studies have looked at RSV reinfections at a community level [25,26,28,29]. Further, the few studies that have considered community settings have investigated either households with an infant and a school going sibling or followed a birth cohort, making their findings less generalizable [19,28,30,31]. Hence, currently there is limited understanding on reinfection incidence and variability among asymptomatic/mild cases, as well as reinfections in older age groups. Additionally the previous studies have utilized partial sequencing of the G and F genes to describe RSV genetic diversity in the context of reinfection [39,40]. However, little is known about genetic changes during reinfection beyond these genes especially in older age groups and in individuals reinfected asymptomatically or with only mild symptoms [31,37,41,42]. Herein, we addressed these gaps by conducting a detailed analysis of RSV infections and reinfections using whole genome sequencing of RSV infections in individuals sampled 1-2 times a week, regardless of symptom status, across diverse age groups during 2023/24 RSV season in coastal Kenya.

## Methods

### Study design and population

We analyzed samples/data collected during the ResViRe_study (**<u>Res</u>**piratory **<u>Vi</u>**rus **<u>Re</u>**infection study) conducted between August 31, 2023, and August 30, 2024. The ResViRe study, run by the KEMRI-Wellcome Trust Research Programme (KWTRP) in Kilifi, Kenya, is a prospective homestead-based respiratory infection surveillance cohort study on the Kenyan coast. The study aims to identify and characterize rates, determinants and transmissibility of respiratory virus reinfections regardless of symptom status. The enrolled homesteads were randomly selected from the northern part of the Kilifi Health and Demographic Surveillance System (KHDSS) area [33]. Homesteads were identified from five administrative locations: Tezo, Ngerenya, Zowerani, Roka, and Matsangoni. For a homestead to be enrolled it had to have at least four members, one of which had to be an infant (aged <12 months), the homestead head consented the homestead to participate, and 75% of the homestead membership consented participation (**Supplementary Material Figure 1**). During the study, weekly home visits were made to enrolled homesteads (n=65, see the definition of a homestead in **Supplementary Material**) and a nasopharyngeal/oropharyngeal (NP/OP) swab was collected from all consenting members. Homesteads were visited twice weekly if a member was found positive for any of the following viruses: influenza A/B, RSVA/B or severe acute respiratory syndrome coronavirus 2 (SARS-CoV-2).

### Laboratory processing

#### RSV detection

Viral RNA was extracted from the 140µL of NP/OP sample using the RNeasy Mini Kit (Qiagen, Hilden, Germany) on the QIAcube^®^ HT instrument as per the manufacturer’s instructions. The RNA extracts were screened for RSVA and RSVB using a multiplex quantitative PCR (qPCR) assay (primers/ probe sequences are provided in **Supplementary Table 1**) [32]. A sample was considered positive if the qPCR cycle threshold (Ct) was ≤ 35.0 and the run’s positive and negative controls were valid.

#### Genome sequencing

Nucleic acid from RSV positive samples was re-extracted using the QIAamp Viral RNA Mini kit (Qiagen, Hilden, Germany) per the manufacturer’s protocol. Complementary DNA (cDNA) synthesis and amplification was done using either SuperScript III One-Step RT-PCR System with Platinum Taq DNA Polymerase (ThermoFisher, Massachusetts, USA) with Group specific primers using a six tiled amplicon strategy[33] or Lunascript reverse transcription kit (New England Biolabs, City, UK). With the latter, cDNA was amplified using Q5® Hot Start High-Fidelity 2x kit (New England Biolabs, City, UK) with RSVA (**Supplementary Table 2**). RSVB sequencing was performed using the ARTIC RSVB amplicon scheme with the Lunascript reverse transcription kit (New England Biolabs, City, UK) and Q5® Hot Start High-Fidelity 2x kit (New England Biolabs, City, UK) protocol [34]. Successful amplification was evaluated by running the PCR products on a 1.5% agarose gel and visualized on a UV transilluminator after staining with Red Safe Nucleic Acid Staining solution (iNtRON Biotechnology Inc., Seoul, South Korea). Samples with successful amplification had their PCR products purified using 1x Agencourt AMPure XP beads and were prepared for whole genome sequencing using Oxford Nanopore Technologies (ONT) platform. Library preparation for ONT sequencing was performed using the Native barcoding sequencing kit (SQK114-NBD96.v14), with native barcodes added for sample multiplexing. The final pooled library was sequenced on the ONT GridION instrument.

### Bioinformatic analyses

#### Genome assembly and lineage assignment

Base-calling and demultiplexing of FAST5 files were performed using the ONT Guppy v6.3.9, and ONT adapters were trimmed with Porechop v0.2.4. The resulting fastq reads were assembled using a reference-based approach using the artic-rsv pipeline with subtype-matched RSV primer schemes and the corresponding (reference genome genbank accession number: OR666573.1 for RSV-A or OR666591.1 for RSV-B). A minimum base quality threshold of Q10 was set.

Consensus genomes were derived by aligning barcode-specific reads to the reference RSV-A /B genomes utilizing an RSVA/RSVB primer scheme. Consensus genomes were refined using raw signals using the with Medaka consensus polishing tool. For positions with low genome read depth (< x10), the ambiguous IUPAC base (N) was assigned. Lineage assignment for consensus genomes used Nextclade v3.10.2 (https://clades.nextstrain.org/) tool following the procedure of Goya and colleagues [35].

#### Phylogenetic analyses

RSV multiple sequence alignments were generated using MAFFT v7.490 checked for misalignments using the program Aliview v1.21. Phylogenetic trees were estimated using the maximum likelihood (ML) approach implemented in IQ-TREE2 v1.6.12 with clustering reliability evaluated by bootstrap resampling (1,000 replicates). These analyses included a subsampled set of global RSV whole genomes from GISAID (https://gisaid.org) collected between January 01 2021 and August 31 2024, for RSVA and January 01 2021 and August 31 2024 for RSVB (details in the **Supplement Material**). ML phylogenetic trees for each RSV Group were linked to origin metadata and visualized using the R package “ggtree” v2.4.1 package. An analysis of amino acid sequence differences between the initial and subsequent RSV infections was performed across the whole genome data set using the Nextclade tool v2.14.1 and visualized using the program SNIPIT v1.4.

### Statistical analyses

Individual demographic and household characteristics – i.e., sex, age, underlying health conditions, housing type, exposure to tobacco smoke within the home, and school attendance – were examined to identify observable differences between individuals who experienced reinfection and those who did not. RSV incidence proportion was calculated as the proportion of participants who experienced at least one RSV infection during the epidemic period. RSV incidence rate was estimated as the number of individual infection episodes divided by the accumulated person-time under observation and expressed per 100 person-years with the person time being the exposure period. Participants who contributed the baseline sample during the study period (n = 28) were excluded from incidence rate calculations, as they did not contribute follow-up time. In addition, individuals who tested RSV-positive at their first sample (n = 5) were excluded from incidence rate estimation, as their true time at risk for a first infection episode could not be determined. For participants with follow-up, person-time at risk was calculated from the date of study entry (first negative sample) until the occurrence of an RSV infection otherwise the end of the study period. The reinfection rate was estimated by dividing the total number of reinfections by the accumulated person-time measured from the first negative sample following the initial episode to the last negative sample preceding a subsequent episode. The working definition of all epidemiological terms used in this analysis is provided in the **Supplement Material**. All data management and analyses were conducted in R version 4.3.0. To protect participant confidentiality, original homestead identifiers were replaced with anonymized codes prior to analysis and presentation.

### Ethics statement

The protocols’ consenting and sample collection process were reviewed and approved by KEMRI Scientific Ethics and Research Unit (SERU), Nairobi Kenya (protocol numbers #3178. Samples were collected following consent from a parent or guardian for participants aged <18-year-olds (with assent for children aged between 13–18-year-old). Individual written informed consent was sought for participants aged >18 years.

## Results

### Participants baseline characteristics

Between August 31, 2023, and August 30, 2024, 672 individuals enrolled in the ResViRe study provided ≥ 1 NP/OP for RSV qPCR testing (median age: 14.25 years (IQR: 4.81-29.65 years), range 3.6 months to 104.4 years at enrollment). The median duration of an individual’s participation was 210 days (IQR: 115-302 days). Participants contributed a median of 35 samples (IQR: 20–47), ranging from 1 to 74 samples. The baseline characteristics of the study participants are provided in **Table 1**. At recruitment, 57.8% (n = 385) of the participants were female, originating from 65 homesteads (107 households). Recruited individuals were distributed across five administrative locations within the KHDSS: Zowerani (n=145), Tezo (n=130), Roka (n=128), Ngerenya (n=140), and Matsangoni (n=130).

**Table 1.**
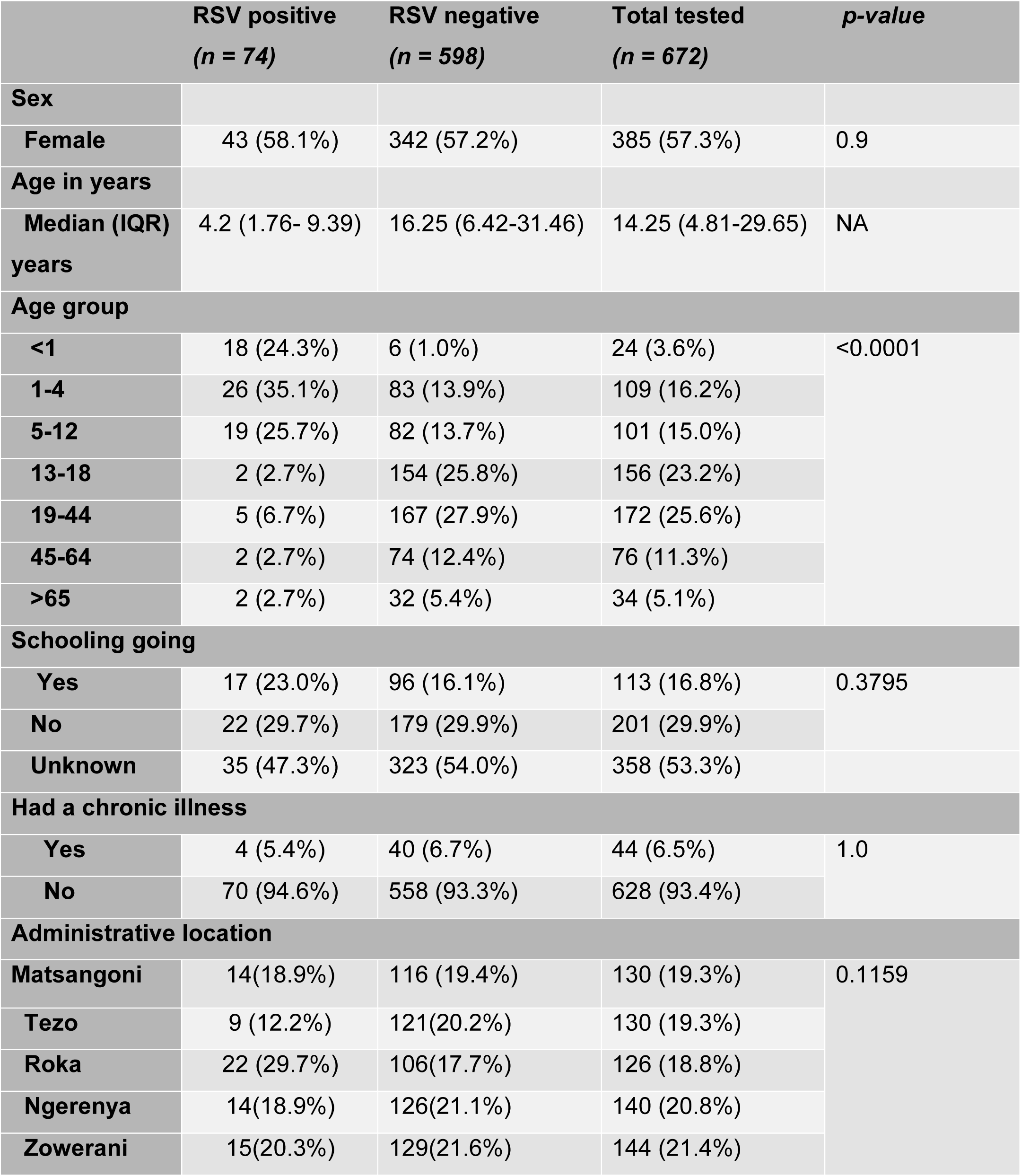
Baseline demographic and epidemiological characteristics of the study participants by RSV status.

|  | RSV positive<br>(n = 74) | RSV negative<br>(n = 598) | Total tested<br>(n = 672) | p-value |
| --- | --- | --- | --- | --- |
| Sex |  |  |  |  |
| Female | 43 (58.1%) | 342 (57.2%) | 385 (57.3%) | 0.9 |
| Age in years |  |  |  |  |
| Median (IQR)<br>years | 4.2 (1.76- 9.39) | 16.25 (6.42-31.46) | 14.25 (4.81-29.65) | NA |
| Age group |  |  |  |  |
| <1 | 18 (24.3%) | 6 (1.0%) | 24 (3.6%) | <0.0001 |
| 1-4 | 26 (35.1%) | 83 (13.9%) | 109 (16.2%) |  |
| 5-12 | 19 (25.7%) | 82 (13.7%) | 101 (15.0%) |  |
| 13-18 | 2 (2.7%) | 154 (25.8%) | 156 (23.2%) |  |
| 19-44 | 5 (6.7%) | 167 (27.9%) | 172 (25.6%) |  |
| 45-64 | 2 (2.7%) | 74 (12.4%) | 76 (11.3%) |  |
| >65 | 2 (2.7%) | 32 (5.4%) | 34 (5.1%) |  |
| Schooling going |  |  |  |  |
| Yes | 17 (23.0%) | 96 (16.1%) | 113 (16.8%) | 0.3795 |
| No | 22 (29.7%) | 179 (29.9%) | 201 (29.9%) |  |
| Unknown | 35 (47.3%) | 323 (54.0%) | 358 (53.3%) |  |
| Had a chronic illness |  |  |  |  |
| Yes | 4 (5.4%) | 40 (6.7%) | 44 (6.5%) | 1.0 |
| No | 70 (94.6%) | 558 (93.3%) | 628 (93.4%) |  |
| Administrative location |  |  |  |  |
| Matsangoni | 14(18.9%) | 116 (19.4%) | 130 (19.3%) | 0.1159 |
| Tezo | 9 (12.2%) | 121(20.2%) | 130 (19.3%) |  |
| Roka | 22 (29.7%) | 106(17.7%) | 126 (18.8%) |  |
| Ngerenya | 14(18.9%) | 126(21.1%) | 140 (20.8%) |  |
| Zowerani | 15(20.3%) | 129(21.6%) | 144 (21.4%) |  |

### RSV positivity trends and incidence rate

During the study period, the 672 individuals provided 22,000 NP/OP swabs for RSV qPCR testing. Twenty-eight participants provided only the baseline sample. Overall, 0.42% (93/22,000) of the swabs tested RSV positive (89 RSVA, 4 RSVB). The positive samples came from 74 (11.0%) participants residing in 37 homesteads (46 households) and were assigned into 80 individual infection episodes (77 RSVA and 3 RSVB). Five participants had their baseline sample positive for RSVA. Infections in the ResVire cohort began on week 47 (November 2023), peaked on weeks 8 and 9 (February 2024), and faded out in week 16 (April 2024) (**Figure 2 & Supplementary Figure 2**). During the months when RSV cases were detected, 563 study participants were sampled and tested at least once. Ninety-nine individuals dropped out of the study before the epidemic started while 14 were recruited after the epidemic ended (week 17, May 2024). An incidence proportion of 11.01% was estimated in the study and an incidence rate of 19.76/100 person-years. Children under-12 months of age had the highest incidence rate (174.0, CI: 103.0-274.0) compared to older age groups (**Supplementary Table 3)**.

Participants who tested RSV positive at least once had a median age of 4.2 years (IQR 1.8-9.4 years) at infection, and 58.1% were females. Thirteen (13) participants had two or more samples testing RSV positive with a median time interval between the positive samples of 6 days, range (2-33 days). Six participants (8.1%) appeared to be reinfected during the outbreak corresponding to a reinfection rate of 13.7/100 person-years. Details of the age, time interval between the infection episodes among the reinfected participants, RSV lineage, and genomic differences between the sequences are given in **Table 2**. Based on baseline health history data, none of the individuals with reinfection reported having a chronic illness. From the associated demographic data, all suspect reinfection participants had housing conditions of mud walls and floor with makuti/iron sheet roof type with only two sharing the household with tobacco users.

**Table 2.** Demographic and virological characteristics of suspected reinfections cases during the ResViRe study Kilifi, Kenya.

| Subject | Age Group(Y) | Sex<sup>#</sup> | Infection episode | Symptom status<sup>£</sup> | Cycle threshold | Episodes intervals (Days) | Lineage | Nucl_diff | Comment |
| --- | --- | --- | --- | --- | --- | --- | --- | --- | --- |
| KELF-01 | 1-5 | M | 1 | S | 28.33 | 28 | A.D.1.11 | 3 | Could be persistent infection |
|  |  |  | 2 | A | 32.41 |  | A.D.1.11 |  |  |
| KELF-02 | 1-5 | F | 1 | A | 34.85 | 35 | A.D.3 | 0 | Could be persistent infection |
|  |  |  | 1 | S | 26.70 |  |  |  |  |
|  |  |  | 1 | A | 32.01 |  | A.D.3 |  |  |
|  |  |  | 2 | A | 28.81 |  | A.D.3 |  |  |
| KELF-03 | 1-5 | M | 1 | A | 23.08 | 52 | A.D.3 | 200 | Confirmed Reinfection |
|  |  |  | 2 | A | 29.33 |  | A.D.2.1 |  |  |
|  |  |  | 2 | A | 30.52 |  | F |  |  |
| KELF-04 | 1-5 | M | 1 | S | 24.29 | 21 | F | 0 | Could be Persistent infection |
|  |  |  | 1 | A | 25.19 |  | A.D.1.11 |  |  |
|  |  |  | 2 | A | 33.63 |  | A.D.1.11 |  |  |
| KELF-05 | 1-5 | M | 1 | A | 33.09 | 30 | A.D.2.1 | 20 | Confirmed reinfection |
|  |  |  | 1 | S | 31.60 |  | A.D.2.1 |  |  |
|  |  |  | 2 | S | 24.60 |  | A. D |  |  |
| KELF-06 | 1-5 | F | 1 | S | 25.13 | 33 | A.D.3 | 78 | Confirmed Reinfection |
|  |  |  | 2 | A | 28.68 |  | A.D.2.1 |  |  |

<sup>£</sup> "A" = asymptomatic, "S" = symptomatic; <sup>#</sup> "F" =Female, "M" = Male

### Clinical presentation and viral load

In forty-one (41) of the 77 RSV-A episodes (53.2%) ≥1 clinical symptoms were reported with 29/77(37.7%) reporting ≥2 symptoms. Among the symptomatic cases 39.0% sought medical attention. This increased with severity of the symptoms – i.e., among those having ≥2 symptoms 55.2% sought heath care services **(Figure 3A)**. The common symptoms recorded among positive cases were: 11 runny nose (26.8%), 9 cough (22.0%), and 6 fever (14.6%) that occurred mainly in individuals <5 years old **(Figure 3B).** The relative infection viral loads were higher for symptomatic cases (i.e., lower qPCR Ct values median 26.51; IQR 23.6-28.3) compared to asymptomatic cases (median Ct 28.9; IQR 25.9-31.9) (**Figure 3B**). Only one of the six suspected reinfection cases showed respiratory symptoms during the second infection episode **Figure B**. Of these individuals, 5/6 had reported symptoms coincident with the visit that collected the first positive sample of the first episode, and the remaining individual reported delayed symptoms in the subsequent visit of the first episode, by which time they had cleared the RSV infection.

### RSV lineages detected in the 2023/24 epidemic

Of the 93 positive NP/OP swabs, RSV sequences (partial or near complete genomes (>80%) were recovered from 76 (87.10%) swabs; 74 RSVA and 2 RSVB (**Figure 1**). These arose from 71 RSVA infection episodes (65 participants) and two RSVB infection episodes (2 participants). Overall, the recovered genome length correlated negatively with the diagnostic qPCR Ct value (**Supplementary Figure 1).** Of the 71 sequenced infection episodes, the RSVA sequences fell into six lineages: A.D (n=1), A.D.1 (n=2), A.D.1.11 (n=21), A.D.2.1 (n=19), A.D.3 (n=30), A.D.5.2 (n=1). The two RSVB sequences fell into lineage B.D.E.1. The monthly temporal detection pattern of the lineages detected in this study is shown in **Figure 3B** while the weekly pattern is shown in **(Supplementary Figure 2).** In Figures 3C and 3D we present the lineage distribution of the RSV sequences collected contemporaneous with our study and deposited on GISAID. For RSVA, A.D.1*, A.D.3*, and A.D.5* were the most prevalent, while for RSVB, B.D.E.1 was the most prevalent.

**Figure 1.**
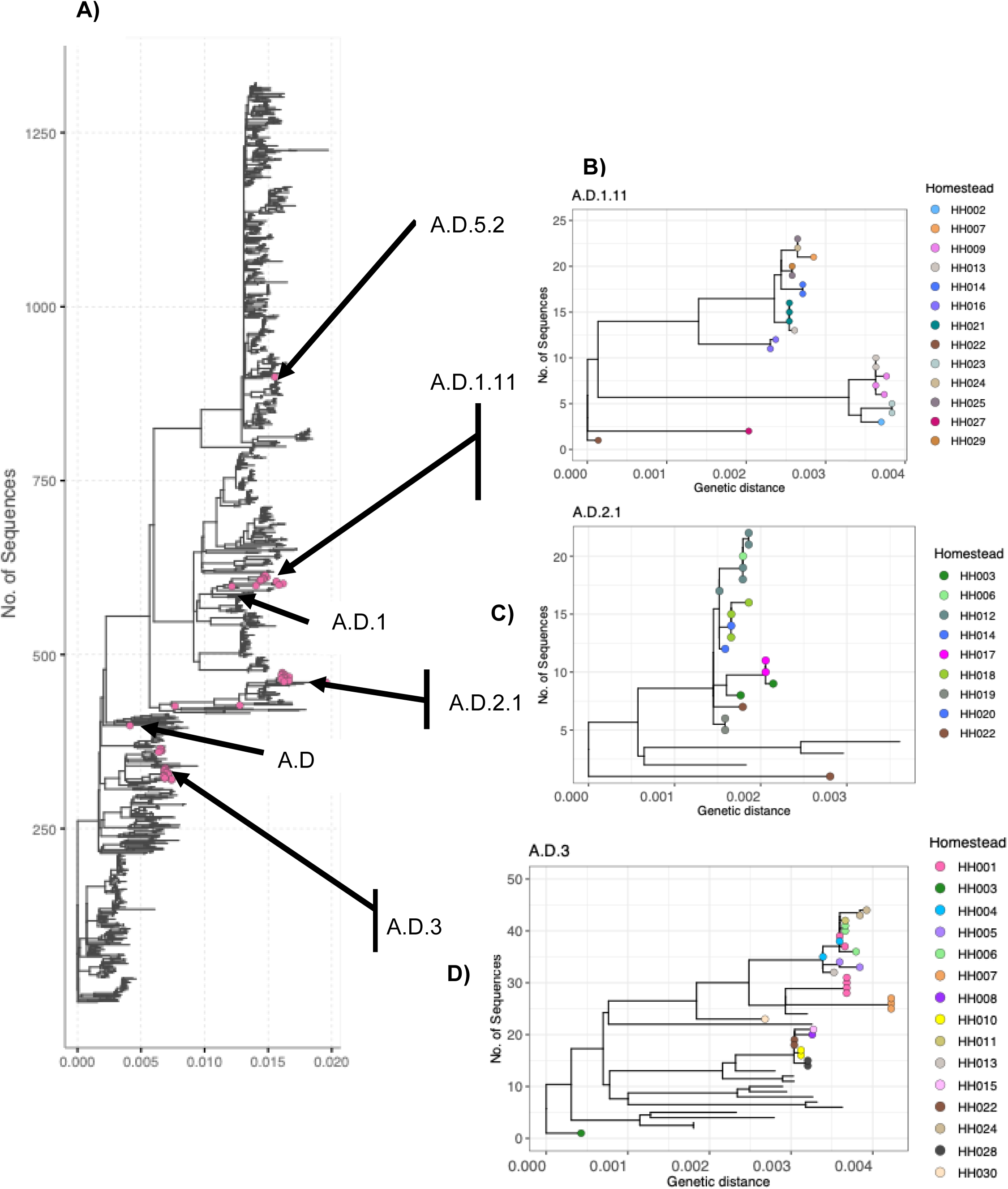
Maximum likelihood phylogenetic tree showing the relationship of the viruses sequenced here a in global context and showing homestead diversity. (A) ML tree showing the clustering of study RSV-A study viruses on the global tree with our study genomes dispersed amongst the contemporary strains circulating in different parts of the world during the study period (study tips coloured red). (B), (C), and (D), ML phylogenetic tree showing the diversity of the A.D.1, A.D.2.1, A.D.3 clades within homesteads. The tip color represents specific homesteads.

**Figure 2.**
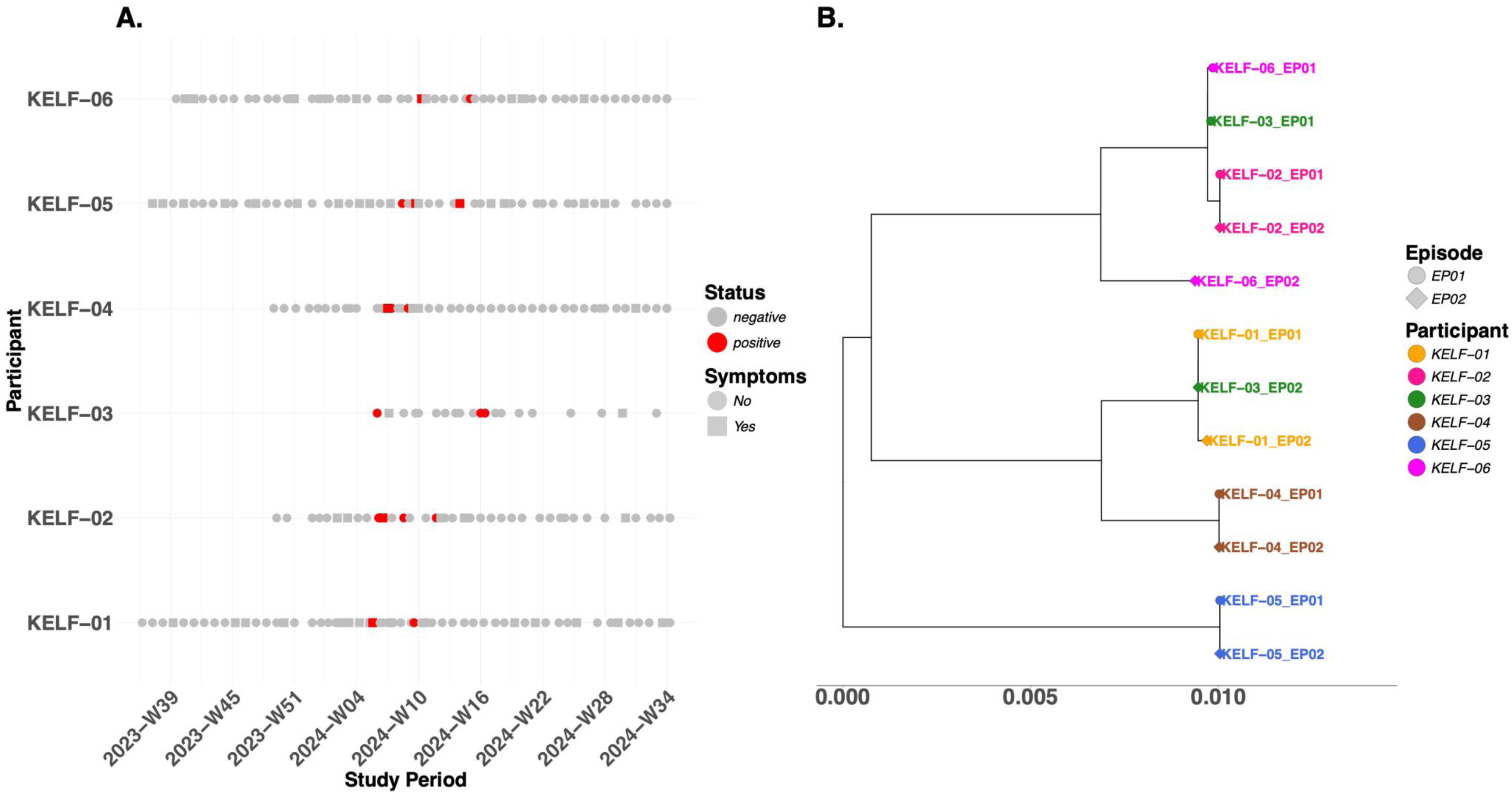
Timelines and evolutionary relationship of suspected reinfections in the ResViRe study. (A) Bubble plot depicting identified reinfections timelines and symptoms status across the study period: grey bubbles indicate negative samples and red bubbles positive samples. The squared shapes indicate symptomatic cases. (B) ML phylogenetic relationships between primary and secondary RSV episodes. The reinfections (EP02) clustered with the primary infection (EP01) cases in majority of the homologous lineage reinfections cases showing little evolution clearly defin

All six suspected reinfections occurred with RSV-A: three involved the same virus lineage, with other three involving a different virus lineage **Table 2**. Among the suspected homologous lineage reinfections, in two the virus pairs were identical (zero nucleotide differences), one had three nucleotide differences. The heterologous lineage reinfections showed 20-200 nucleotide differences (**Table 2**). Comparing amino acid sequences of viruses in reinfection pairs revealed several differences in the G genes **Table 3** and **Supplementary Material Figure 5.** Other amino changes were seen in the F, M2-1, and M2-2 genes **(Supplementary Material Figure 5)** although at lower levels. Other changes were detected in the polymerase genes – in the L, M2-2, and M2-1 genes – although at lower frequency. In addition, participant KLF-06 reinfected by the A.D.3-A.D.2.1 virus lineages showed a loss of an N-glycosylation site at the 273 positions of the G gene.

**Table 3.** G gene amino acid changes detected between the first- and second episode infecting RSV-A viruses identified in the ResViRe study in Kilifi Kenya.

| **EP01 -> EP02** |  |  |  | **EP01 -> EP02** |
| --- | --- | --- | --- | --- |
| **Subject** | **Clades** | **Position** | **Reference** | **Amino Acid change** |
| **KELF-01** | A.D.1 -> A.D.1 | 267<br>285<br>286<br>288 | E<br>S<br>S<br>T | E -> D<br>P -> S<br>P -> S<br>T->A |
| **KELF-03** | A.D.1 -> A.D.3 | 113<br>131<br>142<br>178<br>200<br>225<br>230<br>258<br>269<br>272<br>285<br>286<br>288 | I<br>D<br>L<br>G<br>T<br>V<br>P<br>Q<br>Y<br>P<br>S<br>L<br>S | I -> T<br>D -> V<br>L -> S<br>G -> N<br>T -> A<br>V -> A<br>P -> S<br>Q ->H<br>H -> Y<br>P ->L<br>S -> P<br>L -> P<br>S -> P |
| **KELF-04** | A.D.1 -> A.D.1 | 292 | T | T -> A |
| **KELF-06** | A.D.3 -> A.D.3 | 225<br>269<br>270 | V<br>Y<br>L | V -> A<br>N -> Y<br>P -> L |

### Phylogenetic clustering of the newly sequenced viruses

Phylogenetic analysis of study genomes (coverage >80%, n=74) including global sequences (n=1342) (January 2021-August 2024) revealed a clustering by homestead or household origin within lineages and time of collection (**Figure 4**). The time-resolved ML phylogeny including global sequences (January **2021-August 2024**) suggested that the newly sequenced homesteads’ genomes sequences clustered with those genomes of the same lineage that contemporaneously circulating in other global geographic locations (USA, Europe, Asia, and other parts of Africa) **Supplementary Figure 2B and 2C**.

## Discussion

This study aimed to elucidate the incidence rate, nature and risk factors for RSV infections/reinfections in coastal Kenya population during the 2023/24 epidemic. The study extends our previous investigations by examining reinfections patterns across all age groups regardless of symptom status, inclusion of sensitive molecular diagnostics, and whole genomic characterization of positive cases [26,27,36].

As in previous studies, we report decreasing RSV incidence with increasing age. However, the overall incidence rate of 19.8 infections/100 person-years recorded in this study is lower than that reported by Cohen et al. in South Africa of 47.2/100 person-years [30]. These differences between South Africa and Kenya studies may be due to differences in climate, social economic status, study population and study design (e.g., surveillance months, study households selection, and sample collection frequency). Although our surveillance included persons of all ages, most RSV infections (59.5%) were observed in infants and in young children (aged < 5 years), with the highest incidence rate in under-1-year-olds which is comparable to previous studies [37,38].

During a birth cohort study investigating RSV epidemiology in the first year of life in Kilifi, the estimated incidence rate was ∼3.6 times lower than recorded in this study – i.e., 48.7/100 person-years vs 173.1/100 person-years in the current study [39]. The difference can be attributed to the differences in study design as Nokes et al. [39] sampled/screened the followed infants only when there were ARI symptoms and RSV detection was via antigen detection, whereas the current study collected and tested samples from participants weekly regardless of symptom status and RSV screening was via the more sensitive qPCR.

Of note, we report a relatively low incidence rate of RSV reinfections (13.7 per 100 person-years) consistent with findings reported in other settings globally [28,30,39]. Based on existing literature, sex, household economic status, schooling status, presence of siblings, chronic illness, and household smoke exposure have all been associated with RSV infections [13]. In this study, reinfections were documented only in young children < 2 years. This observation aligns with previous hospital studies where a steady decline in reinfections was observed with advancing age [14,15].

Most (5/6) of the reinfections in the current study presented as asymptomatic with relatively low viral load. Multiple studies suggest that reinfections may occur due to inadequate immune responses or rapid waning of immunity in some individuals [18,26,40]. The existence of RSV reinfections involving the same lineages in the current and past studies within the same epidemic supports the hypothesis of inadequate immune responses in some individuals [18,26]. However, immune responses were not assessed in the current study. Indeed, the low incidence of within epidemic reinfections in this study indicates that approved RSV vaccines and monoclonal antibodies are likely to remain effective within the same season they are used.

The Kilifi 2023/24 RSV epidemic was largely associated with RSVA (95%), specifically the A.D.3, A.D.1.11, A.D.2.1, and A.D.5.2 lineages. Clustering of viruses within the same lineages by homestead/household of origin, confirms that the home environment is a major scene of RSV transmission [19,26]. Three out of the six (3/6) suspected reinfections involved the same virus lineage and occurred within a two-month period. We reported a similar result of short intervals between homologous reinfections in a birth cohort study [26,27]. We cannot exclude some of the suspected reinfections could be persistent infections. Scott et al. further estimated that strain-specific immunity in this population wanes after approximately 6–9 months [27]. Assessment immune responses in this cohort are underway.

In the suspect reinfection cases, most the amino acid variations were seen in the G gene compared to F gene. This supports the notion that the G protein plays a key role in antigenic variability, leading to evasion of immune responses. In addition, genetic variation in the polymerase complex genes L, M2-1 and M2-2 a possibly suggest an ongoing intra-lineage evolution that extends beyond surface glycoproteins. These mutations may reflect subtle adjustments in viral replication and transcription processes that contribute to RSV fitness and enable reinfection.

Our study has some limitations. First, the study focuses on a single RSV epidemic that was dominated by RSV-A which limits the observation of RSV-B reinfection cases. Second, majority of individuals >1 year have likely experienced at least prior RSV infection, and it is challenging to definitively classify subsequent infections as reinfections. This limits the ability to accurately estimate reinfection frequency in older age groups. Third, although the study gives insight to genetic variations and potential risk factors associated with RSV mild reinfections occurring in a community setup, it is based on a small data set which limits potential model analysis to identify specific RSV reinfection risk factors.

In conclusion, this study reinforce that children aged under-2-years-old experience most RSV infection/reinfections in community epidemics regardless of symptom status. The 2023/24 coastal Kenya RSV epidemic was predominated RSV-A; seven lineages co-circulated that showed a close phylogenetic relationship with strains circulating globally. Confirmed reinfections were few, suggesting that the recently introduced immunoprophylactic interventions will be effective at least for one season.

## Supporting information

Supplementary Figure 6B: Global sequences for RSVA

Supplementary Figure 6C: Global sequences for RSVB

## Data Availability

Genomes generates from the study are deposited in Pathoplexus (https://pathoplexus.org/) under the accession numbers PP004EH3D.1 to PP004F1G1.1 for RSVA and accession number PP004F1G1.1 to PP004F1HY.1 for RSVB (see Supplementary Material Table 4).
Epidemiological data and scripts for data analysis are available on the Harvard dataverse https://doi.org/10.7910/DVN/BTIIUZ.

## Acknowledgements

We thank study participants, parents and guardians who consented participation in the studies, members of the Pathogen Epidemiology and Omics (PEO) Group particularly the field team who collected the study samples and metadata and laboratory diagnostics team who processed the samples analysed and presented in this report. We thank Mr. Christopher Nyundo for his assistance in generating the study area map. We thank all genomic data contributors including authors and their originating labs of the sequence data in GISAID that we included in this research. This work is published with permission from director KEMRI.

## Data availability

Genomes deposited in Pathoplexus (https://pathoplexus.org/) under the accession numbers PP_004EH3D.1–PP_ PP_004F1G1.1 for RSV-A and accession number PP_004F1G1.1- PP_004F1HY.1 for RSV-B, and are presented as publicly available in the Pathoplexus database (see **Supplementary Material Table 4).**

Epidemiological data and scripts for data analysis are available on the Harvard dataverse https://doi.org/10.7910/DVN/BTIIUZ.

## References

1. Li Y, Johnson EK, Shi T, et al. National burden estimates of hospitalisations for acute lower respiratory infections due to respiratory syncytial virus in young children in 2019 among 58 countries: a modelling study. Lancet Respir Med [Internet]. 2021 Feb 1 [cited 2024 Nov 12];9(2):175–85. Available from: http://www.thelancet.com/article/S2213260020303222/fulltext

2. Shi T, McAllister DA, O’Brien KL, et al. Global, regional, and national disease burden estimates of acute lower respiratory infections due to respiratory syncytial virus in young children in 2015: a systematic review and modelling study. Lancet [Internet]. 2017 Sep 2 [cited 2024 Nov 12];390(10098):946. Available from: https://pmc.ncbi.nlm.nih.gov/articles/PMC5592248/

3. Liu H, Lu B, Tabor DE, et al. Characterization of human respiratory syncytial virus (RSV) isolated from HIV-exposed-uninfected and HIV-unexposed infants in South Africa during 2015-2017. Influenza Other Respir Viruses [Internet]. 2020 Jul 1 [cited 2024 Nov 7];14(4):403–11. Available from: https://onlinelibrary.wiley.com/doi/full/10.1111/irv.12727

4. Gill CJ, Mwananyanda L, MacLeod WB, et al. Infant deaths from respiratory syncytial virus in Lusaka, Zambia from the ZPRIME study: a 3-year, systematic, post-mortem surveillance project. Lancet Glob Health [Internet]. 2022 Feb 1 [cited 2024 Nov 12];10(2): e269–77. Available from: http://www.thelancet.com/article/S2214109X21005180/fulltext

5. Li Y, Wang X, Blau DM, et al. Global, regional, and national disease burden estimates of acute lower respiratory infections due to respiratory syncytial virus in children younger than 5 years in 2019: a systematic analysis. The Lancet [Internet]. 2022 May 28 [cited 2025 Oct 17];399(10340):2047–64. Available from: https://www.thelancet.com/action/showFullText?pii=S014067362200478 0

6. Korsten K, Adriaenssens N, Coenen S, et al. Burden of respiratory syncytial virus infection in community-dwelling older adults in Europe (RESCEU): An international prospective cohort study. European Respiratory Journal [Internet]. 2021 [cited 2025 Jul 21];57(4). Available from: https://pubmed.ncbi.nlm.nih.gov/33060153/

7. Feldman C, Anderson R. RSV: an overview of infection in adults. Pneumonia 2025 17:1 [Internet]. 2025 Jun 25 [cited 2025 Jul 21];17(1):1–15. Available from: https://pneumonia.biomedcentral.com/articles/10.1186/s41479-025-00165-z

8. Barnes MVC, Openshaw PJM, Thwaites RS. Mucosal Immune Responses to Respiratory Syncytial Virus. Cells 2022, Vol 11, Page 1153 [Internet]. 2022 Mar 29 [cited 2024 Nov 12];11(7):1153. Available from: https://www.mdpi.com/2073-4409/11/7/1153/htm

9. Russell CD, Unger SA, Walton M, Schwarze J. The human immune response to respiratory syncytial virus infection. Clin Microbiol Rev [Internet]. 2017 Apr 1 [cited 2024 Nov 12];30(2):481–502. Available from: https://journals.asm.org/doi/10.1128/cmr.00090-16

10. Alfano F, Bigoni T, Caggiano FP, Papi A. Respiratory Syncytial Virus Infection in Older Adults: An Update. Drugs Aging [Internet]. 2024 Jun 1 [cited 2025 Nov 16];41(6):487. Available from: https://pmc.ncbi.nlm.nih.gov/articles/PMC11193699/

11. Meissner HC. Viral Bronchiolitis in Children. Ingelfinger JR, editor. New England Journal of Medicine [Internet]. 2016 Jan 7 [cited 2025 Nov 16];374(1):62–72. Available from: https://www.nejm.org/doi/pdf/10.1056/NEJMra1413456

12. Rafferty E, Paulden M, Buchan SA, et al. Evaluating the Individual Healthcare Costs and Burden of Disease Associated with RSV Across Age Groups. Pharmacoeconomics [Internet]. 2022 Jun 1 [cited 2025 Oct 27];40(6):633–45. Available from: https://link.springer.com/article/10.1007/s40273-022-01142-w

13. Okiro EA, Ngama M, Bett A, Cane PA, Medley GF, James Nokes D. Factors associated with increased risk of progression to respiratory syncytial virus-associated pneumonia in young Kenyan children*Facteurs associés à l’augmentation des risques de progression vers une pneumonie associée au virus respiratoire syncytial chez les jeunes enfants kenyansFactores asociados con un riesgo aumentado de progresión de la neumonía por virus sincitial respiratorio en niños pequeños en Kenia. Tropical Medicine & International Health [Internet]. 2008 Jul 1 [cited 2025 Apr 18];13(7):914–26. Available from: https://onlinelibrary.wiley.com/doi/full/10.1111/j.1365-3156.2008.02092.x

14. Nduaguba SO, Tran PT, Choi Y, Winterstein AG. Respiratory syncytial virus reinfections among infants and young children in the United States, 2011–2019. PLoS One [Internet]. 2023 Feb 1 [cited 2024 Nov 12];18(2):e0281555. Available from: https://pmc.ncbi.nlm.nih.gov/articles/PMC9934310/

15. Foley DA, Minney-Smith CA, Lee WH, et al. Respiratory Syncytial Virus Reinfections in Children in Western Australia. Viruses [Internet]. 2023 Dec 1 [cited 2024 Nov 12];15(12):2417. Available from: https://www.mdpi.com/1999-4915/15/12/2417/htm

16. Van Drunen Littel-van Den Hurk S, Mapletoft JW, Arsic N, Kovacs-Nolan J. Immunopathology of RSV infection: prospects for developing vaccines without this complication. Rev Med Virol [Internet]. 2007 Jan 1 [cited 2025 Jul 24];17(1):5–34. Available from: /doi/pdf/10.1002/rmv.518

17. Domachowske JB, Rosenberg HF. Respiratory Syncytial Virus Infection: Immune Response, Immunopathogenesis, and Treatment. Clin Microbiol Rev [Internet]. 1999 [cited 2025 Jul 24];12(2):298. Available from: https://pmc.ncbi.nlm.nih.gov/articles/PMC88919/

18. Hall CB, Walsh EE, Long CE, Schnabel KC. Immunity to and Frequency of Reinfection with Respiratory Syncytial Virus. J Infect Dis [Internet]. 1991 Apr 1 [cited 2025 Apr 16];163(4):693–8. Available from: 10.1093/infdis/163.4.693

19. Munywoki PK, Koech DC, Agoti CN, Lewa C, Cane PA, Medley GF, Nokes DJ. The Source of Respiratory Syncytial Virus Infection In Infants: A Household Cohort Study In Rural Kenya. J Infect Dis [Internet]. 2013 Jun 1 [cited 2024 Nov 12];209(11):1685. Available from: https://pmc.ncbi.nlm.nih.gov/articles/PMC4017365/

20. Ohuma EO, Okiro EA, Ochola R, et al. The Natural History of Respiratory Syncytial Virus in a Birth Cohort: The Influence of Age and Previous Infection on Reinfection and Disease. Am J Epidemiol [Internet]. 2012 Nov 1 [cited 2025 Jul 24];176(9):794–802. Available from: 10.1093/aje/kws257

21. Collins PL, Fearns R, Graham BS. Respiratory Syncytial Virus: Virology, Reverse Genetics, and Pathogenesis of Disease. Curr Top Microbiol Immunol [Internet]. 2013 [cited 2025 Sep 19];372:3. Available from: https://pmc.ncbi.nlm.nih.gov/articles/PMC4794264/

22. Goya S, Ruis C, Neher RA, et al. Standardized Phylogenetic Classification of Human Respiratory Syncytial Virus below the Subgroup Level. Emerg Infect Dis [Internet]. 2024 Aug 1 [cited 2025 Oct 21];30(8):1631. Available from: https://pmc.ncbi.nlm.nih.gov/articles/PMC11286072/

23. Bergeron HC, Tripp RA. Immunopathology of RSV: An Updated Review. Viruses [Internet]. 2021 Dec 1 [cited 2025 Oct 24];13(12):2478. Available from: https://pmc.ncbi.nlm.nih.gov/articles/PMC8703574/

24. Taleb SA, Al Thani AA, Al Ansari K, Yassine HM. Human respiratory syncytial virus: pathogenesis, immune responses, and current vaccine approaches. European Journal of Clinical Microbiology and Infectious Diseases [Internet]. 2018 Oct 1 [cited 2025 Oct 24];37(10):1817–27. Available from: https://link.springer.com/article/10.1007/s10096-018-3289-4

25. Okamoto M, Dapat CP, Sandagon AMD, et al. Molecular Characterization of Respiratory Syncytial Virus in Children With Repeated Infections With Subgroup B in the Philippines. J Infect Dis [Internet]. 2018 Aug 24 [cited 2025 Apr 16];218(7):1045–53. Available from: 10.1093/infdis/jiy256

26. Agoti CN, Mwihuri AG, Sande CJ, Onyango CO, Medley GF, Cane PA, Nokes DJ. Genetic Relatedness of Infecting and Reinfecting Respiratory Syncytial Virus Strains Identified in a Birth Cohort From Rural Kenya. J Infect Dis [Internet]. 2012 Nov 15 [cited 2024 Nov 13];206(10):1532. Available from: https://pmc.ncbi.nlm.nih.gov/articles/PMC3475639/

27. Scott PD, Ochola R, Ngama M, Okiro EA, Nokes DJ, Medley GF, Cane PA. Molecular analysis of respiratory syncytial virus reinfections in infants from coastal Kenya. Journal of Infectious Diseases [Internet]. 2006 Jan 1 [cited 2024 Nov 12];193(1):59–67. Available from: 10.1086/498246

28. Munywoki PK, Koech DC, Agoti CN, et al. Influence of age, severity of infection, and co-infection on the duration of respiratory syncytial virus (RSV) shedding. Epidemiol Infect [Internet]. 2015 Mar 25 [cited 2025 Jul 24];143(4):804–12. Available from: https://www.cambridge.org/core/journals/epidemiology-and-infection/article/influence-of-age-severity-of-infection-and-coinfection-on-the-duration-of-respiratory-syncytial-virus-rsv-shedding/34472B89F9319BA11B2C905EA38474BC

29. Yamaguchi M, Sano Y, Dapat IC, et al. High Frequency of Repeated Infections Due to Emerging Genotypes of Human Respiratory Syncytial Viruses among Children during Eight Successive Epidemic Seasons in Japan. J Clin Microbiol [Internet]. 2011 Mar [cited 2025 Apr 17];49(3):1034. Available from: https://pmc.ncbi.nlm.nih.gov/articles/PMC3067727/

30. Cohen C, Kleynhans J, Moyes J, et al. Incidence and transmission of respiratory syncytial virus in urban and rural South Africa, 2017-2018. Nat Commun [Internet]. 2024 Dec 1 [cited 2025 Sep 10];15(1):1–11. Available from: https://www.nature.com/articles/s41467-023-44275-y

31. Nokes DJ, Ngama M, Bett A, et al. Incidence and severity of respiratory syncytial virus pneumonia in rural Kenyan children identified through hospital surveillance. Clin Infect Dis [Internet]. 2009 Nov [cited 2025 Jul 24];49(9):1341. Available from: https://pmc.ncbi.nlm.nih.gov/articles/PMC2762474/

32. Gunson RN, Collins TC, Carman WF. Real-time RT-PCR detection of 12 respiratory viral infections in four triplex reactions. Journal of Clinical Virology [Internet]. 2005 Aug [cited 2024 Nov 13];33(4):341. Available from: https://pmc.ncbi.nlm.nih.gov/articles/PMC7108440/

33. Agoti CN, Otieno JR, Munywoki PK, et al. Local Evolutionary Patterns of Human Respiratory Syncytial Virus Derived from Whole-Genome Sequencing. J Virol [Internet]. 2015 Apr [cited 2025 Jul 25];89(7):3444–54. Available from: https://pubmed.ncbi.nlm.nih.gov/25609811/

34. Maloney DM, Fernandes G, Jasim S, et al. ARTIC RSV amplicon sequencing reveals global RSV genotype dynamics. bioRxiv [Internet]. 2024 Aug 26 [cited 2025 Jul 28];2024.08.23.609324. Available from: https://www.biorxiv.org/content/10.1101/2024.08.23.609324v2

35. Goya S, Ruis C, Neher RA, et al. The unified proposal for classification of human respiratory syncytial virus below the subgroup level. medRxiv [Internet]. 2024 Feb 14 [cited 2025 Jul 25]; 15:2024.02.13.24302237. Available from: https://www.medrxiv.org/content/10.1101/2024.02.13.24302237v1

36. Nokes DJ, Okiro EA, Ngama M, et al. Respiratory Syncytial Virus Infection and Disease in Infants and Young Children Studied from Birth in Kilifi District, Kenya. Clin Infect Dis [Internet]. 2008 Jan 1 [cited 2024 Nov 13];46(1):50. Available from: https://pmc.ncbi.nlm.nih.gov/articles/PMC2358944/

37. Smith M, Kubale J, Kuan G, et al. Respiratory Syncytial Virus Incidence and Severity in a Community-Based Prospective Cohort of Children Aged 0–14 Years. Open Forum Infect Dis [Internet]. 2022 Nov 2 [cited 2025 Nov 20];9(11). Available from: 10.1093/ofid/ofac598

38. Okiro EA, Ngama M, Bett A, Nokes DJ. The Incidence and Clinical Burden of Respiratory Syncytial Virus Disease Identified through Hospital Outpatient Presentations in Kenyan Children. PLoS One [Internet]. 2012 Dec 29 [cited 2025 Dec 8];7(12):e52520. Available from: https://journals.plos.org/plosone/article?id=10.1371/journal.pone.0052520

39. Nokes DJ, Okiro EA, Ngama M, et al. Respiratory Syncytial Virus Epidemiology in a Birth Cohort from Kilifi District, Kenya: Infection during the First Year of Life. J Infect Dis [Internet]. 2004 Nov 15 [cited 2025 Oct 24];190(10):1828–32. Available from: 10.1086/425040

40. Scott PD, Ochola R, Ngama M, Okiro EA, James Nokes D, Medley GF, Cane PA. Molecular Analysis of Respiratory Syncytial Virus Reinfections in Infants from Coastal Kenya. J Infect Dis [Internet]. 2006 Jan 1;193(1):59–67. Available from: 10.1086/498246

