## Supplementary Material: Additional Figures and Tables for "Respiratory syncytial virus infection and reinfection patterns during a community outbreak in Kenya investigated by whole genome sequencing, 2023/2024"

**Respiratory syncytial virus infection and reinfection patterns during a community outbreak on the Kenyan Coast: 2023/24 season**

[Supplementary Table 2: Summary of RSVA whole genome sequencing primers used in this study adapted from Agoti *et al*. [5] with slight modifications. The modified primers are marked with an asterisk.	8](#_Toc218843856)

[Supplementary Figure 10*:* Frequency of amino acid polymorphisms observed in RSVA viruses sequenced in this study (n=74). Panel a. (Top) Structural features of the full-length RSV G protein (amino acids [AA] 1–320); Panel a. (Bottom) Linear plot of individual amino acid variation frequency in full-length RSV A G (red) compared to reference sequence GenBank OR666573.1for the study sequences. Panel b. (Top) Structural features of the full-length RSV F protein (amino acids [AA] 1–574); (Bottom) Linear plot of individual amino acid variation frequency in full-length RSV A F (red) compared to reference sequence GenBank OR666573.1.	25](#_Toc218843863)

### Background

#### RSV interventions

Multiple RSV intervention products are now licensed. These include three vaccines: ABRYSVO® vaccine (Pfizer, USA) that target pregnant mothers, vulnerable adults and older adults (≥ 60 years) [1], AREXVY vaccine (GlaxoSmithKline, UK) that target older adults, mRESVIA vaccine (Moderna, USA) that target vulnerable adults and older adults) [2]. Three monoclonal antibody products are approved for infants entering their first RSV and vulnerable babies <1year in their second RSV season Beyfortus a.k.a Nirsevimab [3] (Astrazeneca (UK) and SANOFI (France)), ENFLONSA™ a.k.a. clesrovimab-cfor (Merck, USA) and SYNAGIS a.k.a palivizumab (AstraZeneca, UK). These products are yet to be rolled out in low- and middle-income countries (LMICs) [5].

### Methods

**Study site and Population**

Participants in the study were recruited from five administrative locations within the Kilifi Health and Demographic surveillance system (KHDSS) area [33**] Figure 1.**

These individuals were distributed across five administrative locations within the KHDSS: Zowerani (n = 145), Tezo (n = 130), Roka (n = 128), Ngerenya (n = 140), and Matsangoni (n = 130 originating from 65 homesteads (107 households).

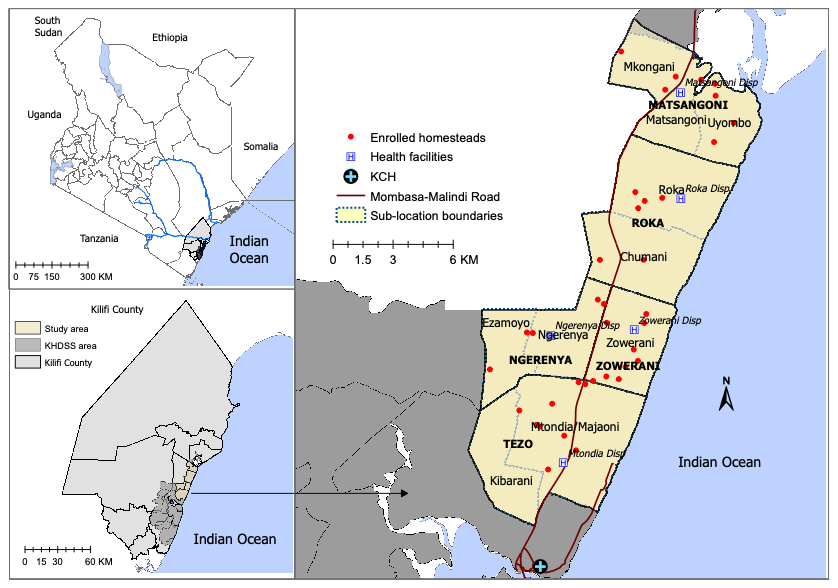

**Supplementary Figure 1:** Study map is showing the study site in the KHDSS and the distribution of the homesteads in the selected locations.

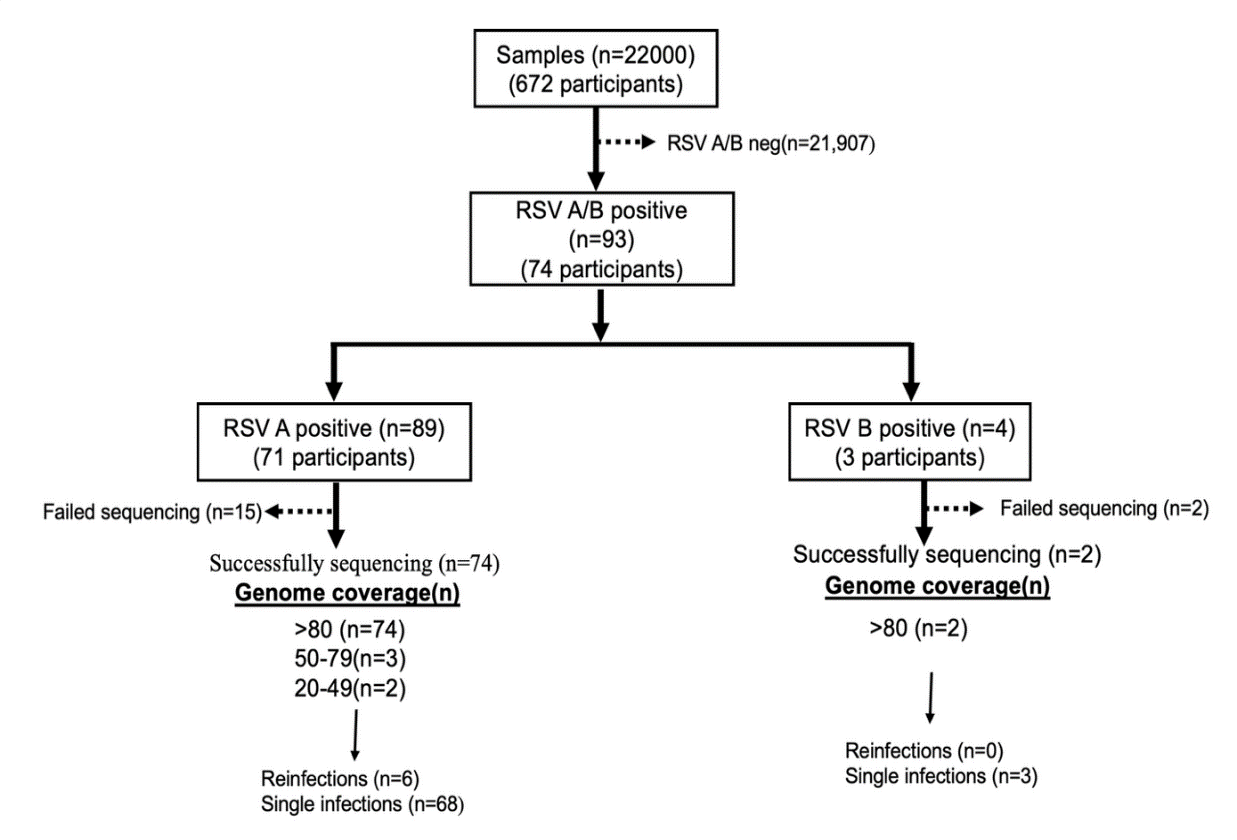

**Supplementary Figure 2:** Flowchart of samples collected during the study. Of the 672 participants, 475 individuals were followed up during the epidemic (November 2023-April 2024).

**Selection of global context data**

We obtained data from Global Initiative on Sharing All Influenza Data (GISAID) (<https://gisaid.org>) collected between January 01, 2021, and August 31, 2024, for RSVA and January 01, 2020, and August 31, 2024, for RSVB. Global data with incomplete date entries or having < 70% coverage were excluded. The global RSV sequence data were organized into a Microsoft Excel database including associated metadata (country of origin, date of isolation, subtype, and genome length. Random subsampling was done using an in-house R script. A set of RSVA (n=1178) and RSVB (n=687) were obtained for further downstream analysis.

#### Prediction of N- and O-Glycosylation Sites

Putative N-glycosylation in the G and F gene (Asn-Xaa-Ser/Thr) in the RSV-A study genomes were predicted using NetNGlyc 1.0 server (threshold ≥0.5) and compared to the reference genome GenBank Accession OR666573.1. For this analysis, the alignment and amino acid position and numbering were based on the reference GenBank accession OR666573.1.

#### RSV Whole Genome Sequencing

RSV-B sequencing was done using Artic RSV-B primer set (<https://github.com/artic-network/artic-rsv>) [6].

#### Supplementary Table 1: Multiplex molecular diagnostic primers used in the study.

| Organism | Target | Sequence (5' to 3') | 5' Modification | 3' Modification |
| --- | --- | --- | --- | --- |
| RSV A | NP | F, AGATCAACTTCTGTCATCCAGCAA  R, TTCTGCACATCATAATTAGGAG  P, CACCATCCAACGGAGCACAGGAGAT | FAM | BHQ1 |
| RSV B | NP | F, AATACAGAAAAATCTAACCAACTTTAC  R, ATATTGCAGCAGTACGCACACA  P, ACACTAGCCATCCTTACTGCGCTTCG | VIC | TAMRA |

#### Supplementary Table 2: Summary of RSVA whole genome sequencing primers used in this study adapted from Agoti *et al*. [5] with slight modifications. The modified primers are marked with an asterisk.

| Target | Primer | Sequence (5′ to 3′) | position | T_m_ (°C) |
| --- | --- | --- | --- | --- |
| RSVA | rsvaS | ACGCGAAAAAATGCGTACAAC | 1 | 57.13 |
|  | rsva50 | GCATGTTATTACAAGTAGTGATATTTGCC | 269 | 57.51 |
|  | rsva52 | TGTGCATGTTATTACAAGTAGTGATATTTG | 266 | 56.96 |
|  | rsva39 | CTTCTCTTAAACCAACCATGGCATC | 2879 | 58.22 |
|  | rsva175 | TTCTCTTAAACCAACCATGGCATCT | 2878 | 58.43 |
| RSVA | rsva86 | AAGAGATGCCATGGTTGGTTTAAGA | 2851 | 58.43 |
|  | rsva117 | ATAAGAGATGCCATGGTTGGTTTAAGA | 2849 | 58.44 |
|  | rsva1644 | CAACTCCATTGTTATTTGCCCC | 5674 | 56.05 |
|  | rsva1688 | CAACTCCATTGTTATTTGCCCCA | 5674 | 57.54 |
| RSVA* | RSV_3_LEFT | CWCTGGGGCAAATAACAATGGAGT | 5615 | 61.0 |
|  | RSV_3_RIGHT | AGGATATTTGTCAGGTAGTATCATTATTTTTGG | 8138 | 62.0 |
| RSVA* | RSV_4_LEFT | ACTGAACTCAACAGCGATGACATC | 7885 | 61.0 |
|  | RSV_4_RIGHT | ATGCTTGATTGAATTTGCTGAGATCTG | 10563 | 60.4 |
| RSVA* | RSV_5_LEFT | AAGAGAACTCAGTGTAGGTAGAATGTTT | 10327 | 60.7 |
|  | RSV_5_RIGHT | TTATATATCCCTCTCCCCAATCYTTTTCAAA | 13007 | 62.2 |
| RSVA* | RSV_6_LEFT | ATTGGGTGTATGCATCTATAGATAACAAG | 12354 | 61.0 |
| RSVA* | RSV_6_RIGHT | TGTATAACAAACTACCTGTGATTTTAATCAG | 14918 | 60.2 |

* Each amplicon was represented by a single primer mapping to the reference sequence, rather than two mapping primers for amplicons 1 and 2 as designed by Agoti et al. (2015).

| Age group (Y) | Person years | Total Positives (n) | Incidence rate/100 | Confidence intervals (95%) |
| --- | --- | --- | --- | --- |
| <1 | 10.4 | 18 | 173.1 | 103.0 – 274.0 |
| 1-4 | 58.43 | 26 | 44.50 | 29.40 – 66.00 |
| 5-12 | 82.99 | 19 | 22.89 | 14.00 – 36.30 |
| 13-18 | 49.0 | 2 | 4.08 | 0.50 – 14.80 |
| 19-44 | 120.0 | 5 | 4.16 | 1.35 – 9.71 |
| 45-64 | 35.5 | 2 | 5.64 | 0.68 – 20.40 |
| >65+ | 17.9 | 2 | 11.17 | 1.35 – 40.40 |

#### Supplementary Table 3: Age group–specific incidence rates of RSV during the 2023/24 RSV epidemic in Kilifi.

**Supplementary Table 4:** RSV-A/B genomes generated in this study and deposited in Pathoplexus. The table includes accession version numbers, RSV subtype, lineage assigned, collection date, and submission identifier.

| **Accession_Version** | **Subtype** | **Lineage** | **Release_Date** | | **Submission_Id** |
| --- | --- | --- | --- | --- | --- |
| PP_004EH3D.1 | A | A.D.1.11 | 2025-12-16 | | Kilifi/RSVA/RVRS/0001_2023_11_30 |
| PP_004EH4B.1 | A | A.D.3 | 2025-12-16 | | Kilifi/RSVA/RVRS/0002_2023_12_19 |
| PP_004EH59.1 | A | A.D.3 | 2025-12-16 | | Kilifi/RSVA/RVRS/0003_2023_12_19 |
| PP_004EH67.1 | A | A.D.2.1 | 2025-12-16 | | Kilifi/RSVA/RVRS/0004_2024_01_15 |
| PP_004EH75.1 | A | A.D.2.1 | 2025-12-16 | | Kilifi/RSVA/RVRS/0005_2024_01_15 |
| PP_004EH83.1 | A | A.D.2.1 | 2025-12-16 | | Kilifi/RSVA/RVRS/0006_2024_01_18 |
| PP_004EH91.1 | A | A.D.2.1 | 2025-12-16 | | Kilifi/RSVA/RVRS/0007_2024_01_19 |
| PP_004EHAZ.1 | A | A.D.3 | 2025-12-16 | | Kilifi/RSVA/RVRS/0008_2024_01_23 |
| PP_004EHBX.1 | A | A.D.1.11 | 2025-12-16 | | Kilifi/RSVA/RVRS/0009_2024_01_24 |
| PP_004EHCV.1 | A | A.D.2.1 | 2025-12-16 | | Kilifi/RSVA/RVRS/0010_2024_01_25 |
| PP_004EHDT.1 | A | A.D.5.2 | 2025-12-16 | | Kilifi/RSVA/RVRS/0011_2024_01_25 |
| PP_004EHER.1 | A | A.D.3 | 2025-12-16 | | Kilifi/RSVA/RVRS/0012_2024_01_26 |
| PP_004EHFP.1 | A | A.D.3 | 2025-12-16 | | Kilifi/RSVA/RVRS/0013_2024_01_26 |
| PP_004EHGM.1 | A | A.D.3 | 2025-12-16 | | Kilifi/RSVA/RVRS/0014_2024_01_26 |
| PP_004EHHJ.1 | A | A.D.3 | 2025-12-16 | | Kilifi/RSVA/RVRS/0015_2024_01_29 |
| PP_004EHJG.1 | A | A.D.3 | 2025-12-16 | | Kilifi/RSVA/RVRS/0016_2024_01_29 |
| PP_004EHKE.1 | A | A.D.2.1 | 2025-12-16 | | Kilifi/RSVA/RVRS/0017_2024_01_29 |
| PP_004EHLC.1 | A | A.D.3 | 2025-12-16 | | Kilifi/RSVA/RVRS/0018_2024_01_29 |
| PP_004EHMA.1 | A | A.D.3 | 2025-12-16 | | Kilifi/RSVA/RVRS/0019_2024_01_31 |
| PP_004EHN8.1 | A | A.D.2.1 | 2025-12-16 | | Kilifi/RSVA/RVRS/0020_2024_02_02 |
| PP_004EHP6.1 | A | A.D.1.11 | 2025-12-16 | | Kilifi/RSVA/RVRS/0021_2024_02_05 |
| PP_004EHQ4.1 | A | A.D.2.1 | 2025-12-16 | | Kilifi/RSVA/RVRS/0022_2024_02_05 |
| PP_004EHR2.1 | A | A.D.1.11 | 2025-12-16 | | Kilifi/RSVA/RVRS/0023_2024_02_06 |
| PP_004EHS0.1 | A | A.D.3 | 2025-12-16 | | Kilifi/RSVA/RVRS/0024_2024_02_06 |
| PP_004EHTY.1 | A | A.D.1.11 | 2025-12-16 | | Kilifi/RSVA/RVRS/0025_2024_02_09 |
| PP_004EHVU.1 | A | A.D.1.11 | 2025-12-16 | | Kilifi/RSVA/RVRS/0027_2024_02_09 |
| PP_004EHWS.1 | A | A.D.3 | 2025-12-16 | | Kilifi/RSVA/RVRS/0028_2024_02_12 |
| PP_004EHXQ.1 | A | A.D.2.1 | 2025-12-16 | | Kilifi/RSVA/RVRS/0029_2024_02_12 |
| PP_004EHYN.1 | A | A.D.1.11 | 2025-12-16 | | Kilifi/RSVA/RVRS/0030_2024_02_13 |
| PP_004EHZL.1 | A | A.D.1.11 | 2025-12-16 | | Kilifi/RSVA/RVRS/0031_2024_02_15 |
| PP_004EJ0J.1 | A | A.D.3 | 2025-12-16 | | Kilifi/RSVA/RVRS/0032_2024_02_16 |
| PP_004EJ1G.1 | A | A.D.3 | 2025-12-16 | | Kilifi/RSVA/RVRS/0033_2024_02_20 |
| PP_004EJ2E.1 | A | A.D.3 | 2025-12-16 | | Kilifi/RSVA/RVRS/0034_2024_02_20 |
| PP_004EJ3C.1 | A | A.D.1.11 | 2025-12-16 | | Kilifi/RSVA/RVRS/0035_2024_02_22 |
| PP_004EJ4A.1 | A | A.D.2.1 | 2025-12-16 | | Kilifi/RSVA/RVRS/0036_2024_02_23 |
| PP_004EJ58.1 | A | A.D.3 | 2025-12-16 | | Kilifi/RSVA/RVRS/0037_2024_02_24 |
| PP_004EJ66.1 | A | A.D.3 | 2025-12-16 | | Kilifi/RSVA/RVRS/0038_2024_02_24 |
| PP_004EJ74.1 | A | A.D.1.11 | 2025-12-16 | | Kilifi/RSVA/RVRS/0039_2024_02_24 |
| PP_004EJ82.1 | A | A.D.3 | 2025-12-16 | | Kilifi/RSVA/RVRS/0040_2024_02_26 |
| PP_004EJ90.1 | A | A.D.3 | 2025-12-16 | | Kilifi/RSVA/RVRS/0041_2024_02_27 |
| PP_004EJAY.1 | A | A.D.3 | 2025-12-16 | | Kilifi/RSVA/RVRS/0042_2024_02_28 |
| PP_004EJBW.1 | A | A.D.1.11 | 2025-12-16 | | Kilifi/RSVA/RVRS/0043_2024_02_28 |
| PP_004EJCU.1 | A | A.D.2.1 | 2025-12-16 | | Kilifi/RSVA/RVRS/0044_2024_02_28 |
| PP_004EJDS.1 | A | A.D.1.11 | 2025-12-16 | | Kilifi/RSVA/RVRS/0045_2024_02_29 |
| PP_004EJEQ.1 | A | A.D.3 | 2025-12-16 | | Kilifi/RSVA/RVRS/0046_2024_03_01 |
| PP_004EJFN.1 | A | A.D.3 | 2025-12-16 | | Kilifi/RSVA/RVRS/0047_2024_03_01 |
| PP_004EJGL.1 | A | A.D.3 | 2025-12-16 | | Kilifi/RSVA/RVRS/0048_2024_03_01 |
| PP_004EJHH.1 | A | A.D.2.1 | 2025-12-16 | | Kilifi/RSVA/RVRS/0049_2024_03_01 |
| PP_004EJJF.1 | A | A.D.1.11 | 2025-12-16 | | Kilifi/RSVA/RVRS/0050_2024_03_04 |
| PP_004EJKD.1 | A | A.D.3 | 2025-12-16 | | Kilifi/RSVA/RVRS/0051_2024_03_04 |
| PP_004EJLB.1 | A | A.D.3 | 2025-12-16 | | Kilifi/RSVA/RVRS/0052_2024_03_05 |
| PP_004EJM9.1 | A | A.D.1.11 | 2025-12-16 | | Kilifi/RSVA/RVRS/0053_2024_03_05 |
| PP_004EJN7.1 | A | A.D.1.11 | 2025-12-16 | | Kilifi/RSVA/RVRS/0054_2024_03_05 |
| PP_004EJP5.1 | A | A.D.1.11 | 2025-12-16 | | Kilifi/RSVA/RVRS/0055_2024_03_05 |
| PP_004EJQ3.1 | A | A.D.1.11 | 2025-12-16 | | Kilifi/RSVA/RVRS/0056_2024_03_06 |
| PP_004EJR1.1 | A | A.D.1.11 | 2025-12-16 | | Kilifi/RSVA/RVRS/0057_2024_03_08 |
| PP_004EJSZ.1 | A | A.D.3 | 2025-12-16 | | Kilifi/RSVA/RVRS/0058_2024_03_13 |
| PP_004EJTX.1 | A | A.D.1 | 2025-12-16 | | Kilifi/RSVA/RVRS/0059_2024_03_15 |
| PP_004EJUV.1 | A | A.D.3 | 2025-12-16 | | Kilifi/RSVA/RVRS/0060_2024_03_18 |
| PP_004EJVT.1 | A | A.D.3 | 2025-12-16 | | Kilifi/RSVA/RVRS/0061_2024_03_18 |
| PP_004EJWR.1 | A | A.D.3 | 2025-12-16 | | Kilifi/RSVA/RVRS/0062_2024_03_23 |
| PP_004EJXP.1 | A | A.D.2.1 | 2025-12-16 | | Kilifi/RSVA/RVRS/0063_2024_03_26 |
| PP_004EJYM.1 | A | A.D.5.2 | 2025-12-16 | | Kilifi/RSVA/RVRS/0064_2024_03_27 |
| PP_004EJZK.1 | A | A.D.2.1 | 2025-12-16 | | Kilifi/RSVA/RVRS/0065_2024_04_03 |
| PP_004EK0H.1 | A | A.D.3 | 2025-12-16 | | Kilifi/RSVA/RVRS/0066_2024_04_08 |
| PP_004EK1F.1 | A | A.D.2.1 | 2025-12-16 | | Kilifi/RSVA/RVRS/0067_2024_04_08 |
| PP_004EK2D.1 | A | A.D | 2025-12-16 | | Kilifi/RSVA/RVRS/0068_2024_04_08 |
| PP_004EK3B.1 | A | A.D.3 | 2025-12-16 | | Kilifi/RSVA/RVRS/0069_2024_04_15 |
| PP_004EK49.1 | A | A.D.2.1 | 2025-12-16 | | Kilifi/RSVA/RVRS/0070_2024_04_22 |
| PP_004GQ8S.1 | A | A.D.3 | 2026-01-07 | | Kilifi/RSVA/RVRS/0072_2023_12_19 |
| PP_004GQ9Q.1 | A | A.D.1.11 | 2026-01-07 | | Kilifi/RSVA/RVRS/0073_2024_01_23 |
| PP_004GQAN.1 | A | A.D.1.11 | 2026-01-07 | | Kilifi/RSVA/RVRS/0074_2024_01_26 |
| PP_004GQBL.1 | A | A.D.2.1 | 2026-01-07 | | Kilifi/RSVA/RVRS/0075_2024_01_29 |
| PP_004GQCJ.1 | A | A.D.1.11 | 2026-01-07 | | Kilifi/RSVA/RVRS/0076_2024_01_30 |
| PP_004GQDG.1 | A | A.D.3 | 2026-01-07 | | Kilifi/RSVA/RVRS/0077_2024_02_05 |
| PP_004GQEE.1 | A | A.D.1.11 | 2026-01-07 | | Kilifi/RSVA/RVRS/0078_2024_02_09 |
| PP_004GQFC.1 | A | A.D.1 | 2026-01-07 | | Kilifi/RSVA/RVRS/0079_2024_02_13 |
| PP_004GQGA.1 | A | A.D.3 | 2026-01-07 | | Kilifi/RSVA/RVRS/0080_2024_02_24 |
| PP_004GQH7.1 | A | A.D.3 | 2026-01-07 | | Kilifi/RSVA/RVRS/0081_2024_02_27 |
| PP_004GQJ5.1 | A | A.D.1.11 | 2026-01-07 | | Kilifi/RSVA/RVRS/0082_2024_02_28 |
| PP_004GQK3.1 | A | A.D.2.1 | 2026-01-07 | | Kilifi/RSVA/RVRS/0083_2024_02_29 |
| PP_004GQL1.1 | A | A.D.3 | 2026-01-07 | | Kilifi/RSVA/RVRS/0084_2024_03_01 |
| PP_004GQMZ.1 | A | A.D.3 | 2026-01-07 | Kilifi/RSVA/RVRS/0085_2024_03_05 | |
| PP_004GQNX.1 | A | A.D.3 | 2026-01-07 | Kilifi/RSVA/RVRS/0086_2024_03_08 | |
| PP_004GQPV.1 | A | A.D.2.1 | 2026-01-07 | Kilifi/RSVA/RVRS/0087_2024_03_09 | |
| PP_004GQQT.1 | A | A.D.2.1 | 2026-01-07 | Kilifi/RSVA/RVRS/0088_2024_03_18 | |
| PP_004GQRR.1 | A | A.D.3 | 2026-01-07 | Kilifi/RSVA/RVRS/0089_2024_04_04 | |
| PP_004F1G1.1 | B | B.D.E.1 | 2025-12-22 | Kilifi/RSVB/RVRS/0001_2024_03_01 | |
| PP_004F1HY.1 | B | B.D.E.1 | 2025-12-22 | Kilifi/RSVB/RVRS/0002_2024_03_01 | |

**Supplementary Figure 3*:*** RSV positivity trends and genetic diversity observed during the study period. A. Combined bar and line graph showing weekly detection of RSV-A/B viruses during the study period. The bars show positive proportion of RSV subgroup (RSV-A grey28 and RSV-B black) while the line graph (red) shows the total samples tested collected during the study period. B. Weekly occurrence of the 7 RSV-A/B clades occurring during the study period from August 2023/24.

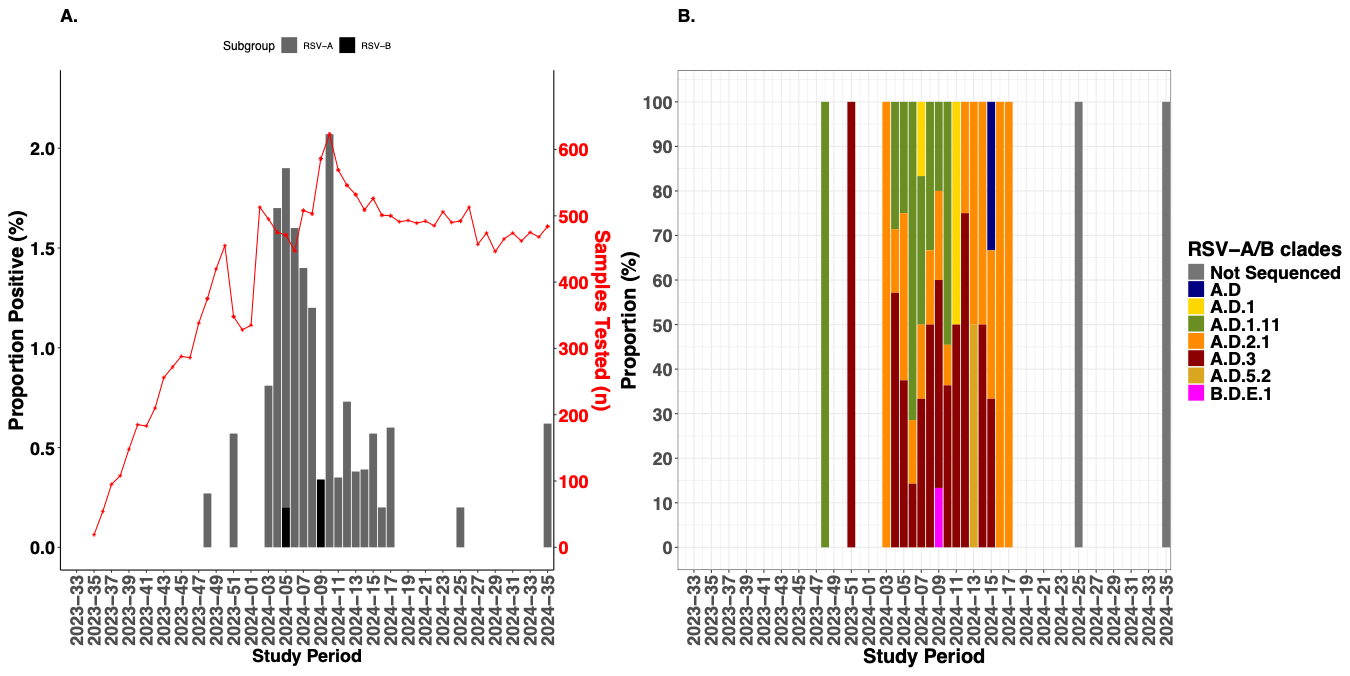

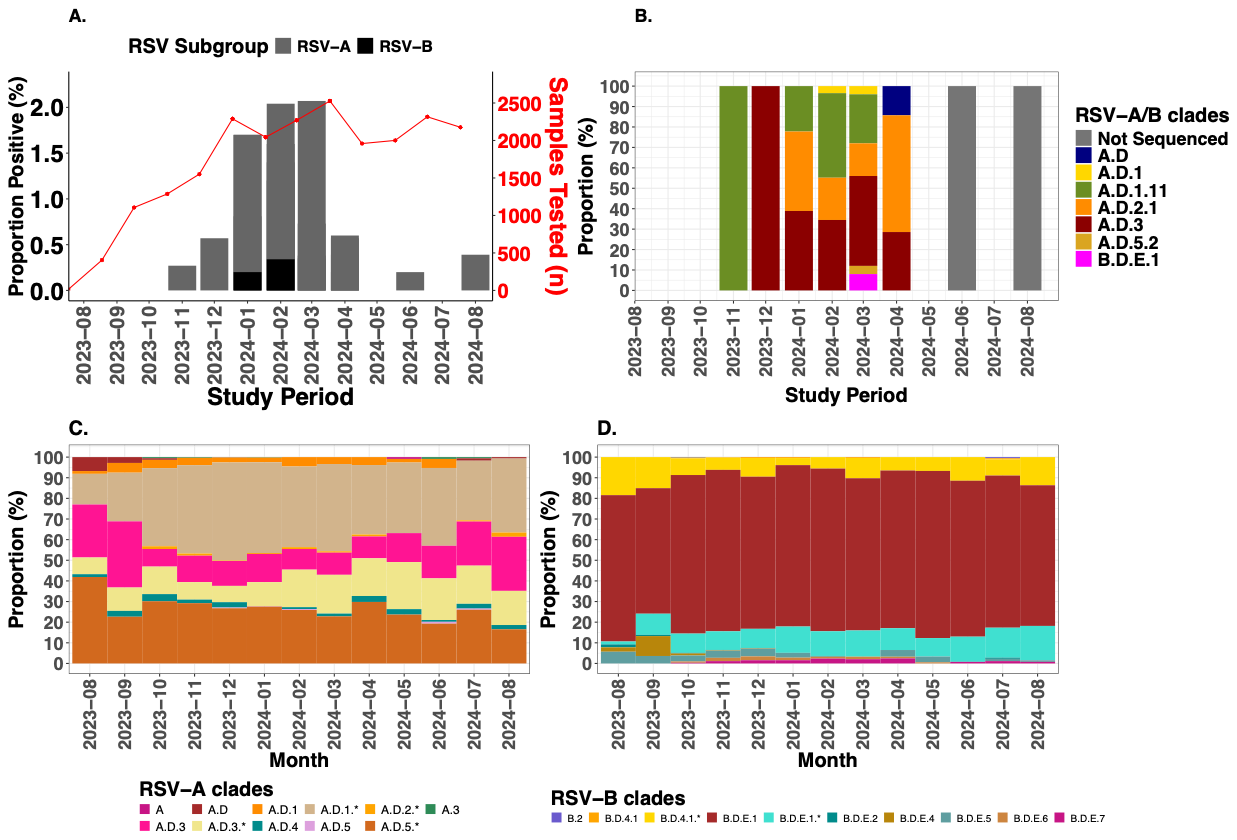

**Supplementary Figure 4:** Temporal patterns and genetic diversity of RSV sequences occurring during the study period in the study region and globally. (A) Combined bar and line graph depicting monthly detection frequency of RSV-A and RSV-B viruses during the study period: The bar graph shows RSV detection by sub-type while the line graph (red) shows the total number collected during the study period. (B) Monthly occurrence of the RSV-A/B clades occurring during the study period from August 2023/24. (C) Diversity of the RSV-A clades circulating globally during the study period 1^st^ August 2023 – 31^st^ August 2024. (D) Diversity of the RSV-B clades circulating globally during the study period 1^st^ August 2023 – 31^st^ August 2024.

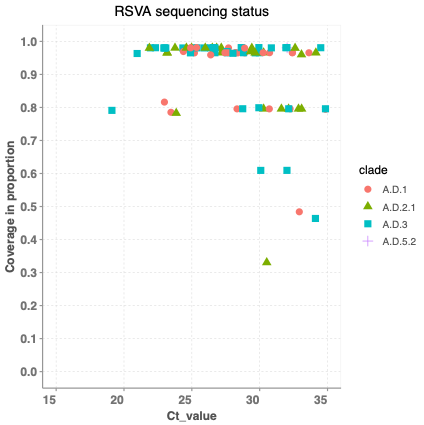

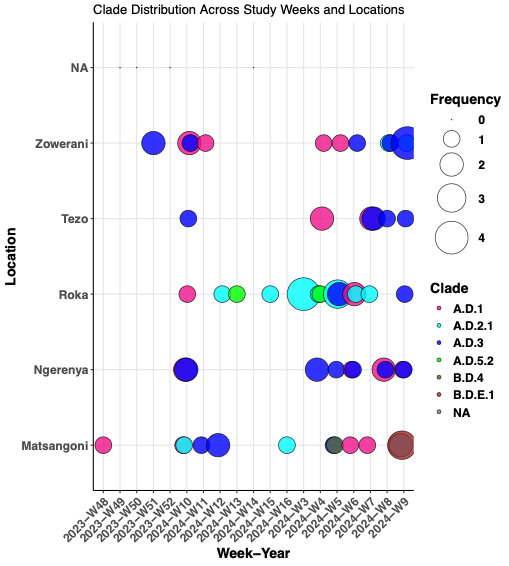

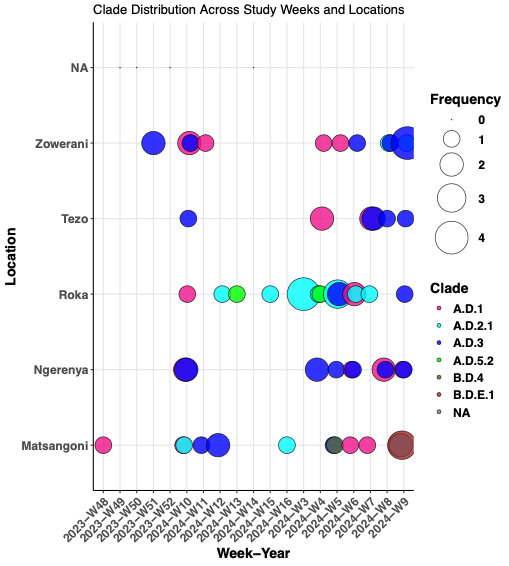

A.

B.

#### Supplementary Figure 5: (A) Relationship between obtained genome coverage and cycle threshold (Ct) values. (B) Bubble plot illustrating the distribution of RSV clades detected at the different study locations.

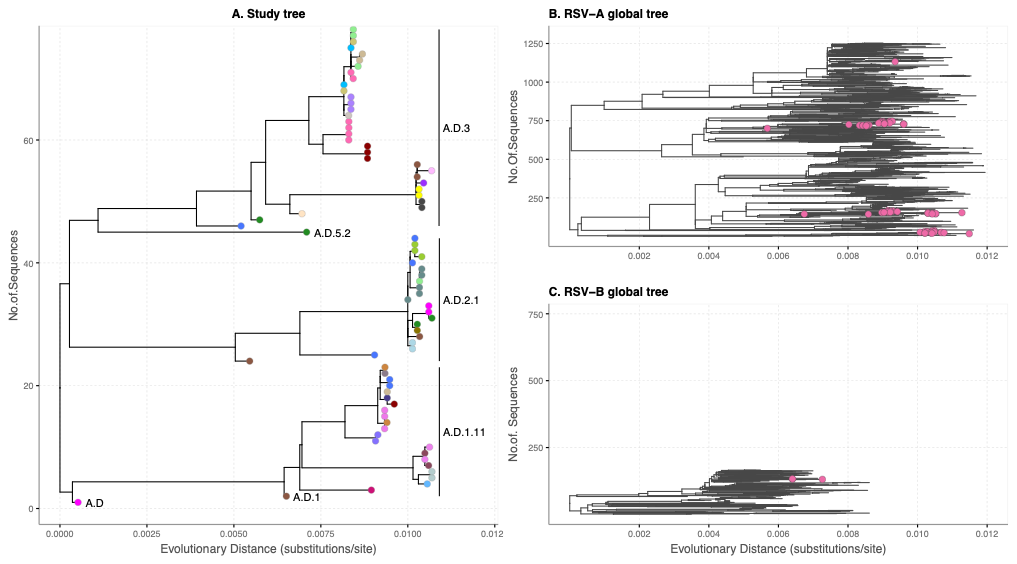

#### Supplementary Figure 6: (A) Maximum Llkelihood divergence phylogenetic trees, (B) clustering of the study sequences coloured based on study homesteads, and (C) RSV-A clustering of study sequences relative to the global contemporaneous RSV-A sequences (n=1178).

Supplementary Figure 7: Root-to-tip regression of genetic divergence versus sampling date for RSV-A (A) and RSV-B (B) sequences. Outlier sequences (highlighted in red) were excluded from further analyses to improve clock-likeness.

A.

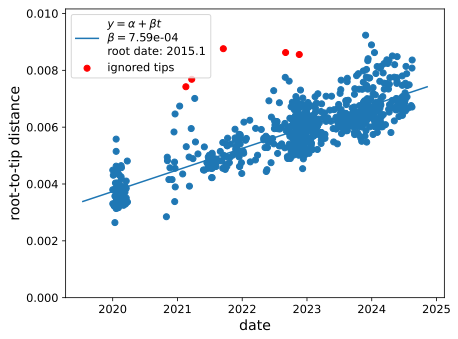

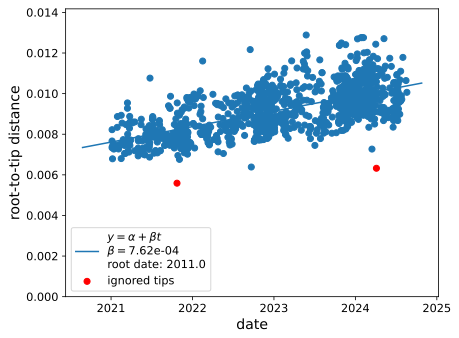

B.

#### Supplementary Figure 8: F gene amino acid polymorphisms observed in reinfection cases in relative between the reference sequences OR666573.1.

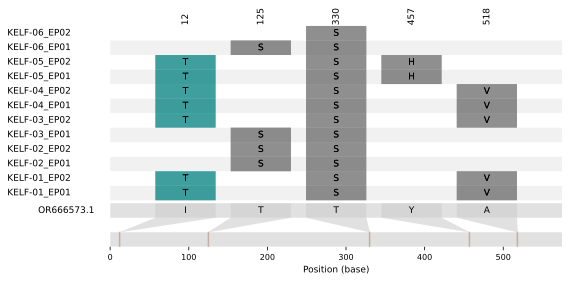

#### Supplementary Figure 9: Amino acid polymorphisms observed in reinfection cases a. L, b. M2-1 and c. M2-2 genes shown relative to the reference sequence OR666573.1.

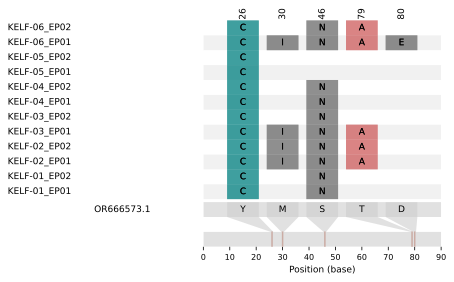

**a.**

**b.**

**c.**

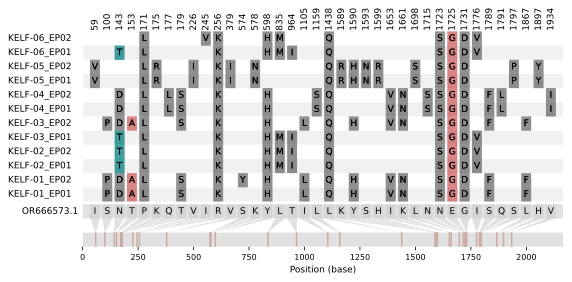

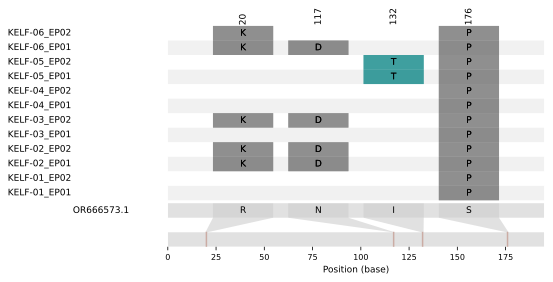

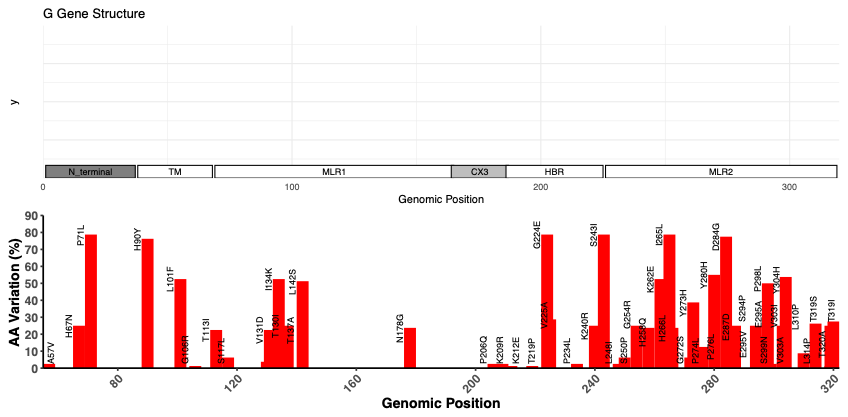

**b.**

**a.**

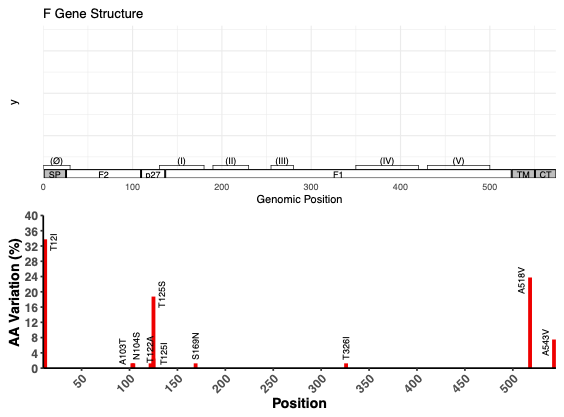

#### Supplementary Figure 10*:* Frequency of amino acid polymorphisms observed in RSVA viruses sequenced in this study (n=74). Panel a. (Top) Structural features of the full-length RSV G protein (amino acids [AA] 1–320); Panel a. (Bottom) Linear plot of individual amino acid variation frequency in full-length RSV A G (red) compared to reference sequence GenBank OR666573.1for the study sequences. Panel b. (Top) Structural features of the full-length RSV F protein (amino acids [AA] 1–574); (Bottom) Linear plot of individual amino acid variation frequency in full-length RSV A F (red) compared to reference sequence GenBank OR666573.1.

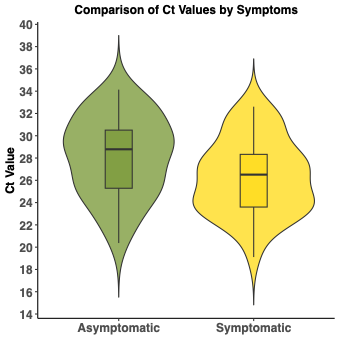

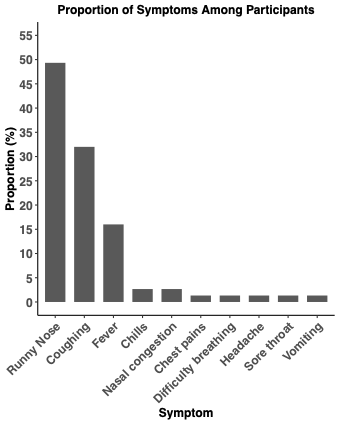

A.

B.

#### Supplementary Figure 11: Respiratory symptoms associated with episodes of RSV infection in a rural Kenyan prospective household study. (A) Frequency distribution of the most common symptoms reported among symptomatic episodes. (B) Comparison of viral load between asymptomatic and symptomatic episodes.
